# Pubertal Growth Spurt Characteristics of Vietnamese Urban Schoolchildren: Population and Individual Level Analysis Using SITAR

**DOI:** 10.64898/2026.07.26.26358986

**Authors:** Nhan T. Ho, Quyet V. Nguyen, Anh Q. Dao, Chi T. L. Tran, An N. Pham

**Affiliations:** Research Management Department, Vinmec International Hospital, Hanoi, Vietnam; College of Health Sciences, VinUniversity, Hanoi, Vietnam; Health Check Department, Vinmec Times City International Hospital, Vinmec International Hospital, Hanoi, Vietnam; Pediatric Department, Vinmec Central Park International Hospital, Vinmec International Hospital, Hanoi, Vietnam; Pediatric Department, Vinmec Haiphong International Hospital, Vinmec International Hospital, Haiphong, Vietnam; Pediatric Department, Vinmec Times City International Hospital, Vinmec International Hospital, Hanoi, Vietnam

**Author notes:** **Corresponding authors:** Nhan T. Ho, Research Management Department, Vinmec International Hospital, Hanoi, Vietnam, and College of Health Sciences, VinUniversity, Hanoi, Vietnam.

**Keywords:** puberty, growth spurt, SITAR, peak height velocity, adolescent growth, Vietnam, BMI, secular trend

## Abstract

Pubertal growth spurt characteristics vary substantially across populations, but no study has characterized these parameters in Vietnamese children using the SITAR (SuperImposition by Translation And Rotation) framework, despite Vietnam’s rapid ongoing nutritional transition. We fitted sex specific SITAR models to longitudinal height data from 15,491 children (9,047 boys, 6,444 girls, 2018 to 2025) to estimate population and individual age at peak height velocity (APHV) and peak height velocity (PHV), and used multivariable linear mixed effects and logistic regression models to identify prepubertal predictors of pubertal timing and intensity. Population mean APHV was 12.2 years in boys and 9.2 years in girls, with PHV of 9.3 and 7.0 cm/year, respectively. This is earlier than previously reported Korean, Colombian, Ethiopian and US cohorts. Individual variability in pubertal timing was nearly twice as large in girls as in boys. After multivariable adjustment, higher baseline BMI-for-age z-score independently predicted earlier pubertal timing in both sexes, whereas height-for-age z-score and prepubertal height velocity showed opposite direction associations (earlier timing in boys but later timing in girls). A significant secular trend toward earlier puberty was observed in girls (∼1 year per decade) but not in boys. Logistic models predicting early pubertal timing achieved acceptable discrimination in boys (AUC=0.76) and modest discrimination in girls (AUC=0.70). This first SITAR-based characterization study reveals earlier pubertal timing in these Vietnamese urban schoolchildren than comparator populations, pronounced sex-specific associations between prepubertal BMI status and height status, and a marked secular trend in earlier puberty in girls warranting further surveillance.

## Introduction

The adolescent growth spurt is one of the most conspicuous milestones of human development, marking the transition from a stable prepubertal growth rate to a period of rapid, transient acceleration in stature that concludes near-final adult height. Two parameters derived from longitudinal height velocity curves capture this process including age at peak height velocity (APHV) (the age at which growth is fastest), and peak height velocity (PHV) itself. Together these are increasingly recognized as validated anthropometric biomarkers of an individual’s chronobiological and maturational status, applicable both to individual clinical assessment and to population-level growth surveillance (1). The SITAR (SuperImposition by Translation And Rotation) model has become a widely adopted method for estimating these parameters (2), because it simultaneously yields a population mean growth curve and child-specific random effects describing each individual’s size, timing, and intensity of the pubertal growth spurt relative to that population mean. Notably, the growth spurt marks the period during which children reach roughly 90% of their final height, following a broadly uniform pattern of growth and skeletal maturation across sexes, despite substantial variation in its timing and magnitude between individuals (3).

An expanding body of SITAR-based literature has documented considerable heterogeneity in pubertal growth spurt parameters across populations. Korean children reach peak height velocity at approximately 12.5 years in boys and earlier in girls, with a comparatively short spurt duration (4). Chinese cohorts, both nationally and in city-specific samples such as Zhongshan, show similar East Asian patterns with urban-rural and sex differences in timing (5,6). Greek children show an adolescent growth spurt preceded by a gradual hormonal rise in IGF-1 and estradiol before the appearance of physical pubertal signs (7). In Latin America, Colombian, Peruvian, and Brazilian cohorts each report distinct pubertal timing profiles, with altitude exerting a modest but detectable influence on peak height velocity and final height in some, though not all, of these populations (8–11). Population-level chronobiological framing of PHV as a biomarker further underscores that timing and magnitude of the growth spurt reflect population-specific, and possibly sex-specific, growth strategies rather than a single universal pattern (1). Despite this rich comparative literature spanning East Asia, Latin America, Europe, and East Africa (12), no study has yet applied the SITAR framework to characterize population-level pubertal growth spurt parameters in a Vietnamese or, more broadly, mainland Southeast Asian cohort. This is a substantial gap given the region’s rapid, ongoing nutritional transition.

Characterizing pubertal growth spurt timing and intensity also carries downstream clinical relevance. Earlier pubertal onset, smaller spurt intensity, and shorter spurt duration have each been linked to lower final height and higher risk of overweight and obesity in late adolescence in Chinese children (5). Large-scale genome-wide association analyses indicate that distinct longitudinal pubertal height-growth patterns carry shared heritability with diverse adult health outcomes, with no single pattern appearing uniformly “optimal” (13). The timing of the growth spurt has further been linked to ocular growth and myopia onset in Singaporean schoolchildren (14), while pubertal changes in body composition (fat mass and fat-free mass) covary with the timing of peak height velocity in Korean adolescents (15). In clinically vulnerable populations, such as adolescents with perinatal HIV infection, pubertal growth spurts are frequently delayed or blunted (16), illustrating that spurt characterization has utility well beyond height prediction alone. At the same time, evidence from healthy adolescents suggests that the timing of individual pubertal milestones does not, by itself, meaningfully alter expected adult height (17), reinforcing the need to examine timing, intensity, and their anthropometric correlates jointly rather than any single parameter in isolation.

Vietnam is undergoing a rapid nutritional transition, with rising childhood overweight and obesity prevalence coexisting alongside residual stunting (18), a pattern that plausibly shapes pubertal growth spurt characteristics in ways not yet described. No prior study has applied the SITAR framework to a Vietnamese pediatric cohort to jointly estimate population-level APHV and PHV, characterize individual-level heterogeneity in pubertal timing and intensity, and examine their association with prepubertal anthropometric status. In this study, we used eight years of longitudinal school-based anthropometric surveillance from the three major cities in Vietnam (Hanoi, Hochiminh, Haiphong) to fit sex-specific SITAR models, derive population and individual pubertal growth spurt parameters, benchmark these against previously published cohorts (4,8–12,19), and identify prepubertal predictors of earlier pubertal timing and altered growth spurt intensity in Vietnamese urban schoolchildren.

## Methods

### Study Population and Data Source

This study analyzed deidentified longitudinal annual school health check data from a private school system of Hanoi, Ho Chi Minh City, and Haiphong, Vietnam from 2018 to 2025.

### Anthropometric Classification

Height-for-age and BMI-for-age were classified using WHO growth references: the WHO Child Growth Standards for children ≤5 years (20) and the WHO 2007 growth reference for children and adolescents aged 5 to 19 years (21). Continuous height-for-age z-scores (HAZ) and BMI-for-age z-scores (BAZ) were computed using the standard WHO method, scaling each child’s deviation from the age- and sex-specific median by the locally appropriate standard deviation on either side of the median. Height-for-age status was categorized as stunting (HAZ < -2), normal, or extreme tallness (height > +3 SD). BMI-for-age status was categorized as thinness (BAZ < -2), normal, overweight (BAZ > +1), or obesity (BAZ > +2), consistent with WHO cut-offs (20,21).

### SITAR Model for Pubertal Growth Spurt Characterization

Sex-specific SITAR (SuperImposition by Translation And Rotation) growth curve models (2) were fitted to longitudinal height measurements to characterize the population and individual pubertal growth spurt. SITAR models a common population growth curve (represented by a natural cubic spline) together with three child-specific random effects that translate and rotate the population curve to fit each child’s individual trajectory: size (a, a vertical shift reflecting overall stature), tempo (b, a horizontal shift in timing, where negative values indicate earlier pubertal maturation relative to the population mean), and velocity/intensity (c, a scaling of the age axis, where positive values indicate a slower or smaller-magnitude growth spurt) (2).

Given the biological difference in pubertal timing between sexes, sex-specific fitting windows were used to ensure each model’s age range fully captured the pubertal growth spurt while remaining within the range of observed data. Boys were modeled over ages 6 to 18 years and girls over ages 6 to 16 years, reflecting girls’ earlier attainment of near-final height. Children required a minimum of four height measurements within the respective fitting window to be included, to ensure stable estimation of individual random effects. Cubic spline degrees of freedom for the population mean curve were set separately for each sex (5 for boys, 4 for girls) to accommodate the different widths of the two fitting windows. Models were fitted using the R sitar package (2), which implements the underlying nonlinear mixed-effects model via the nlme framework (22).

Population mean age at peak height velocity (APHV) and peak height velocity (PHV) were derived from each sex-specific model’s fitted mean velocity curve as the age and magnitude, respectively, at which the first derivative of the fitted height curve was maximal. Individual-level estimated APHV was calculated as the population mean APHV plus each child’s estimated tempo random effect (b). Ninety-five percent confidence intervals (95%CI) for the population mean APHV and PHV were calculated using the normal approximation (mean ± 1.96 × standard error of the mean), where the standard error was derived from the empirical standard deviation of individual-level APHV and PHV estimates across all children divided by the square root of the sample size.

### Prepubertal Anthropometric Predictors

To examine associations between prepubertal growth and subsequent pubertal timing and intensity, sex-specific prepubertal age cutoffs were defined a priori as ≤11 years for boys and ≤9 years for girls, chosen to precede the population mean APHV in each sex with an adequate safety margin. For each child, the following prepubertal predictors were derived from visits occurring at or before the sex-specific cutoff: baseline height, HAZ, and BAZ at the earliest prepubertal visit, short-term prepubertal height velocity (cm/year), calculated from the two earliest prepubertal visits separated by 0.4 to 3 years, and the annual rate of change in BAZ (ΔBAZ/year), estimated as the slope of a child-level linear regression of BAZ on age across all available prepubertal visits with non-missing BAZ (minimum two visits). Prepubertal BMI trajectory group (Normal, overweight/obesity-related, or thinness-related) was derived by comparing BMI category at the earliest and latest available prepubertal visits, with a child classified as overweight/obesity-related or thinness-related if either endpoint visit fell into the corresponding category.

### Statistical Analysis

Cohort characteristics are compared between sexes using the Kruskal-Wallis test for continuous variables and the chi-squared test for categorical variables.

Population mean APHV and PHV, with 95% CI, were compared descriptively against previously published SITAR-derived estimates from Korean (4), Ethiopian (12), Colombian (9) and US (19) cohorts, selected because each study applied the similar SITAR modeling framework (2) to longitudinal height data in a comparable age range, allowing direct comparison of model-derived pubertal timing and intensity parameters across populations.

Associations between prepubertal BMI category and SITAR timing (b), intensity (c), and individual estimated APHV were assessed within each sex using the Kruskal-Wallis test, given the non-normal distribution of SITAR random effects. Trajectories of HAZ and BAZ across the SITAR fitting window were visualized by BMI category and sex using generalized additive model (GAM) smoothers with cubic regression splines and 95% CI using via the mgcv package (23).

Multivariable linear mixed-effects models (LMEs) were fitted separately by sex to identify independent predictors of pubertal timing (b) and intensity (c), each including standardized (z-scored) baseline BAZ, baseline HAZ, prepubertal height velocity, ΔBAZ/year, age at baseline, and birth year as fixed effects, with a random intercept for city to account for geographic clustering. Models were fitted using the lme4 and lmerTest packages (24,25) with maximum likelihood estimation and the BOBYQA optimizer. Standardizing continuous predictors allowed direct comparison of relative effect sizes within each outcome and sex. Marginal and conditional R² and the intraclass correlation coefficient (ICC) for the city random effect were computed for each fitted model.

To evaluate predictors of early pubertal timing as a binary outcome, sex-specific multivariable logistic regression models were fitted with early puberty (SITAR b < 0, i.e. earlier than the sex-specific population mean) as the outcome and the same standardized prepubertal predictors plus city (categorical, with Hanoi as reference) as covariates, with backward-forward stepwise selection by Akaike Information Criterion (AIC). Model discrimination was quantified using the area under the receiver operating characteristic curve (AUC) with 95% CI computed via the pROC package (26).

Geographic variation in individual estimated APHV across the three study cities was visualized descriptively by sex. Secular trends in individual estimated APHV by birth year were assessed using simple linear regression fitted separately by sex, with birth year as the sole predictor.

All analyses were conducted in R version 4.5.1 (27). Two-sided p-values <0.05 were considered statistically significant.

## Results

### Cohort characteristics

Of 97,030 children with valid anthropometric records after quality control, 20,082 (10,848 boys, 9,234 girls) had sufficient longitudinal height data to fit individual SITAR curves (≥4 visits within the fitting window, boys 6 to 18 years, girls 6 to 16 years). After linking SITAR parameters to prepubertal baseline anthropometry, the final analytic cohort comprised 15,491 children (9,047 boys, 6,444 girls) (**Table 1**). Boys entered the prepubertal assessment window at an older median age than girls (7.4 vs 6.8 years), consistent with the later sex-specific cutoff applied for boys. Median baseline height was 127.0 cm in boys and 121.1 cm in girls. Prepubertal BMI status differed markedly by sex in that obesity prevalence was substantially higher in boys than girls at baseline (26.5% vs 7.7%), while normal BMI was more common in girls (70.1% vs 49.7%, all sex comparisons p<0.001) (**Table 1**). Distribution across the three study cities (Hanoi, Ho Chi Minh City, Haiphong) did not differ significantly by sex (p=0.089).

**Table 1.**
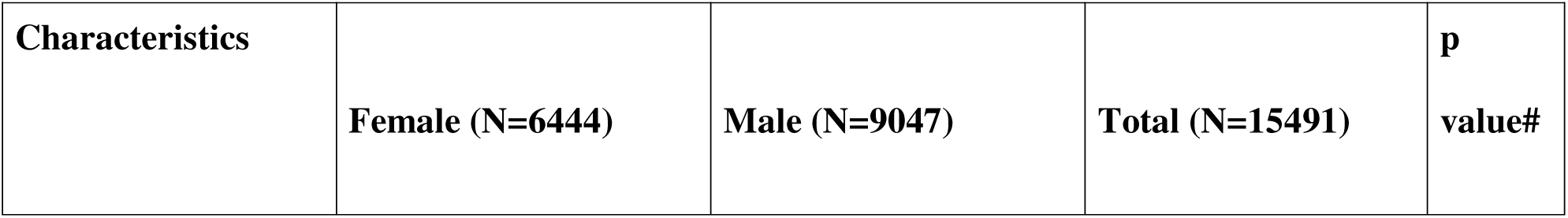
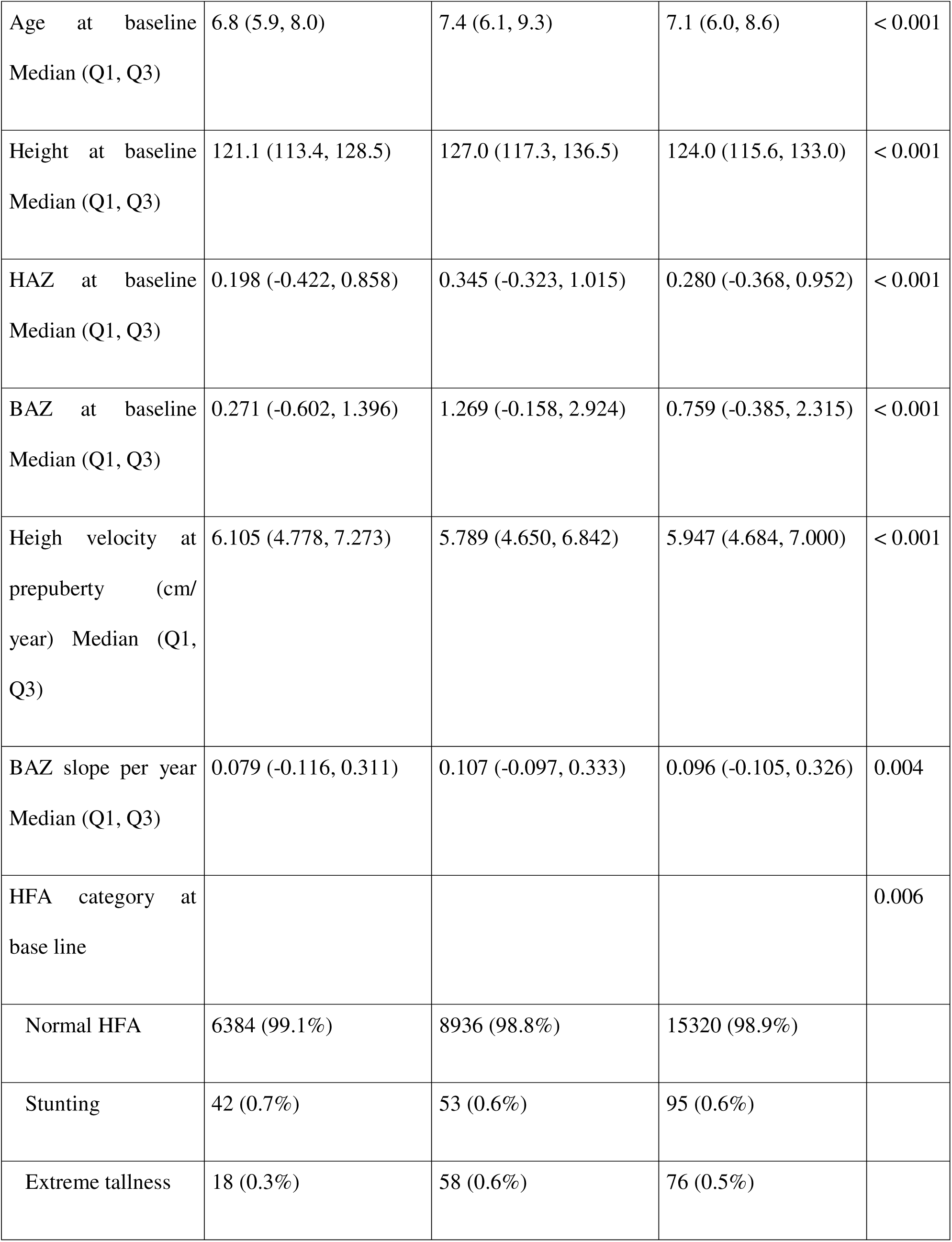
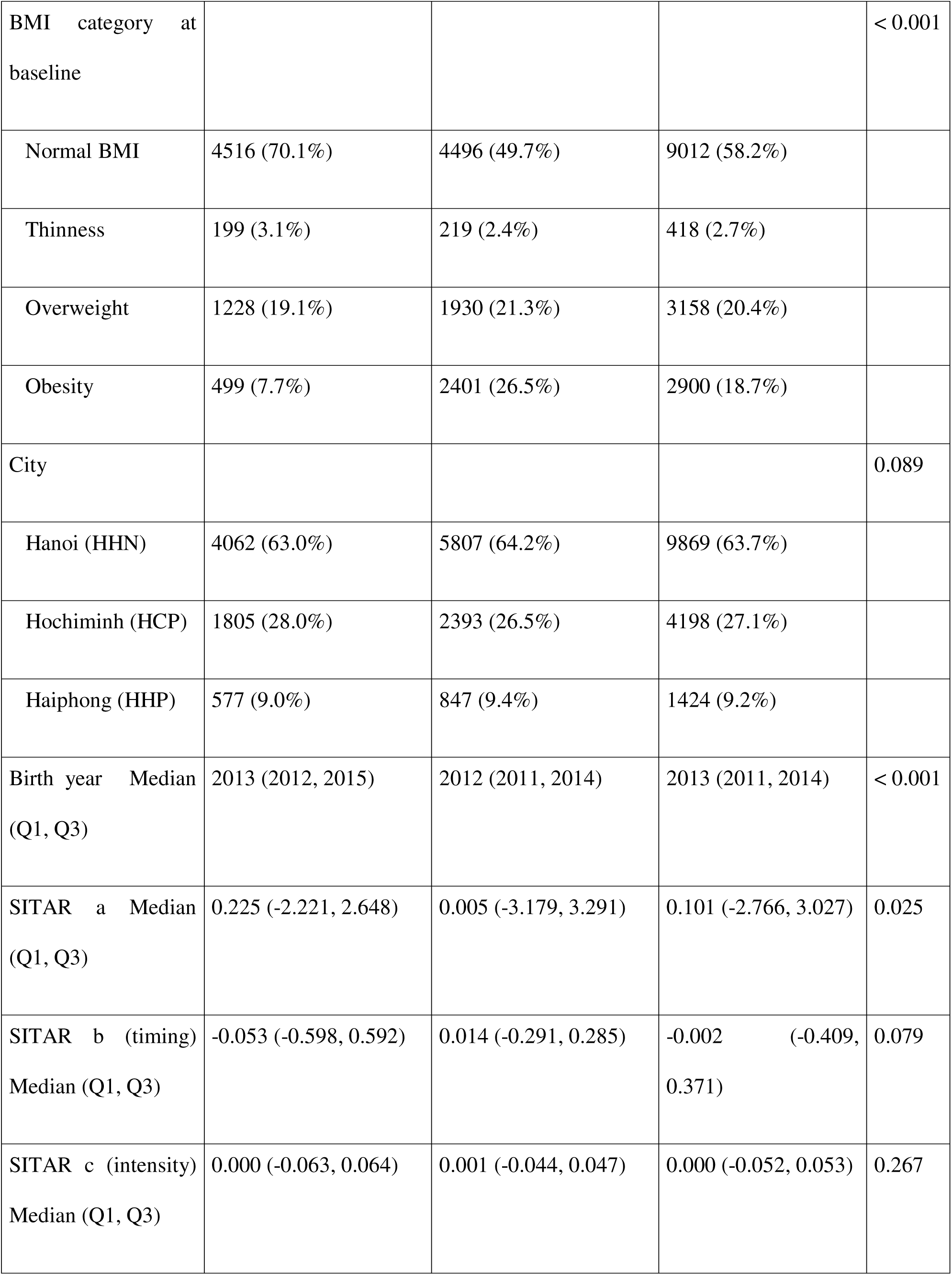
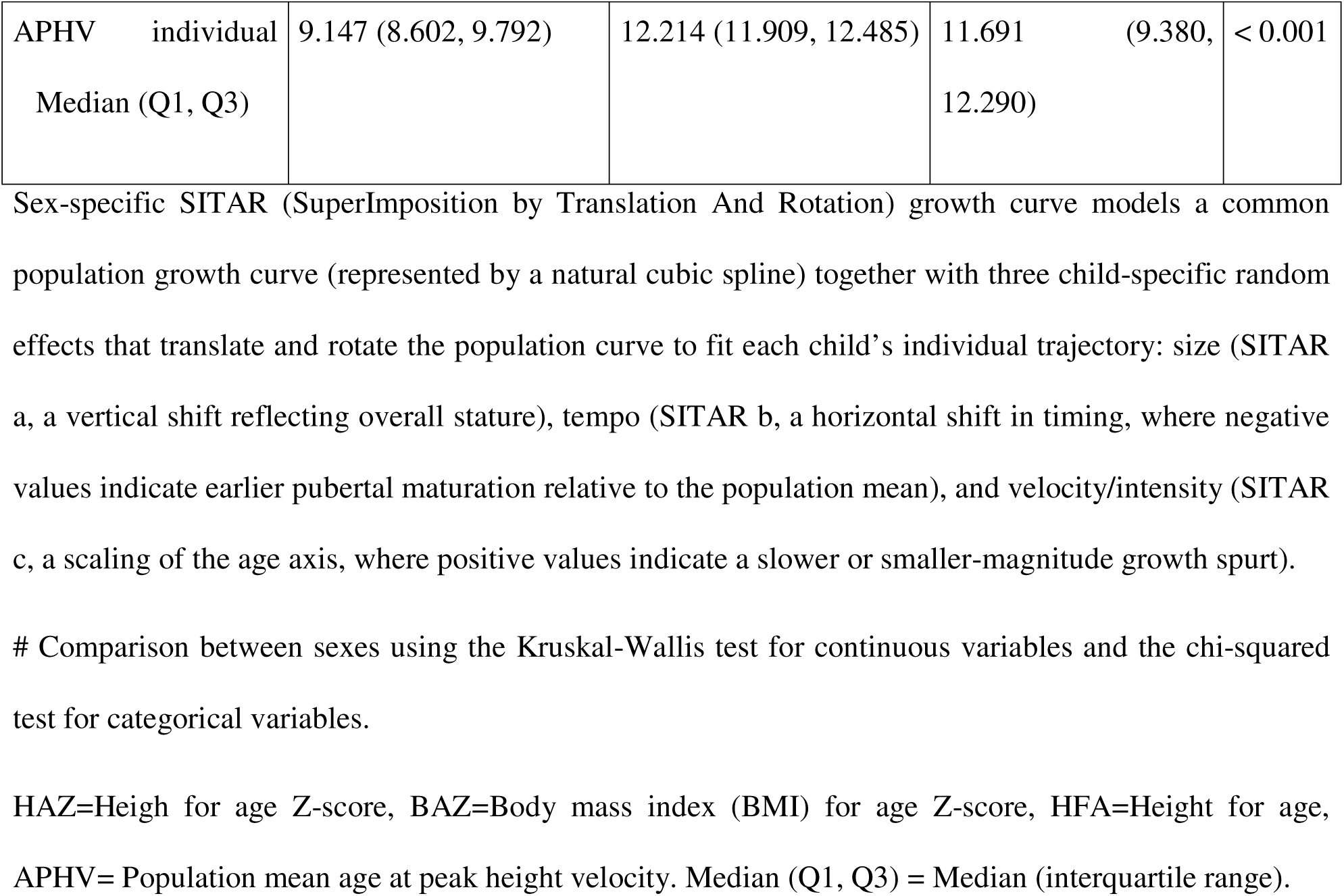
Cohort characteristics by sex.

### Population-level pubertal growth spurt parameters

Sex-specific SITAR models fitted to the full longitudinal height data yielded population mean age at peak height velocity (APHV) of 12.2 (95% CI = 12.19, 12.21) years in boys and 9.2 (95%CI = 9.18, 9.22) years in girls, with corresponding peak height velocity (PHV) of 9.3 cm/year (95%CI= 9.33, 9.36 cm/year) and 7.0 cm/year (95%CI= 6.98, 7.02 cm/year), respectively (**Figure 1a**, **Table 2**). Compared with previously published SITAR-derived estimates, Vietnamese children in this cohort reached peak height velocity earlier than Korean (4), Colombian (9), Ethiopian (12) and US (19) children of the same sex, with the largest difference observed relative to the Ethiopian cohort (2.0 years earlier in boys, 3.3 years earlier in girls) (**Table 2**). Peak height velocity was correspondingly higher in the Vietnamese cohort than in Korean, Colombian and Ethiopian population for males.

**Figure 1.**
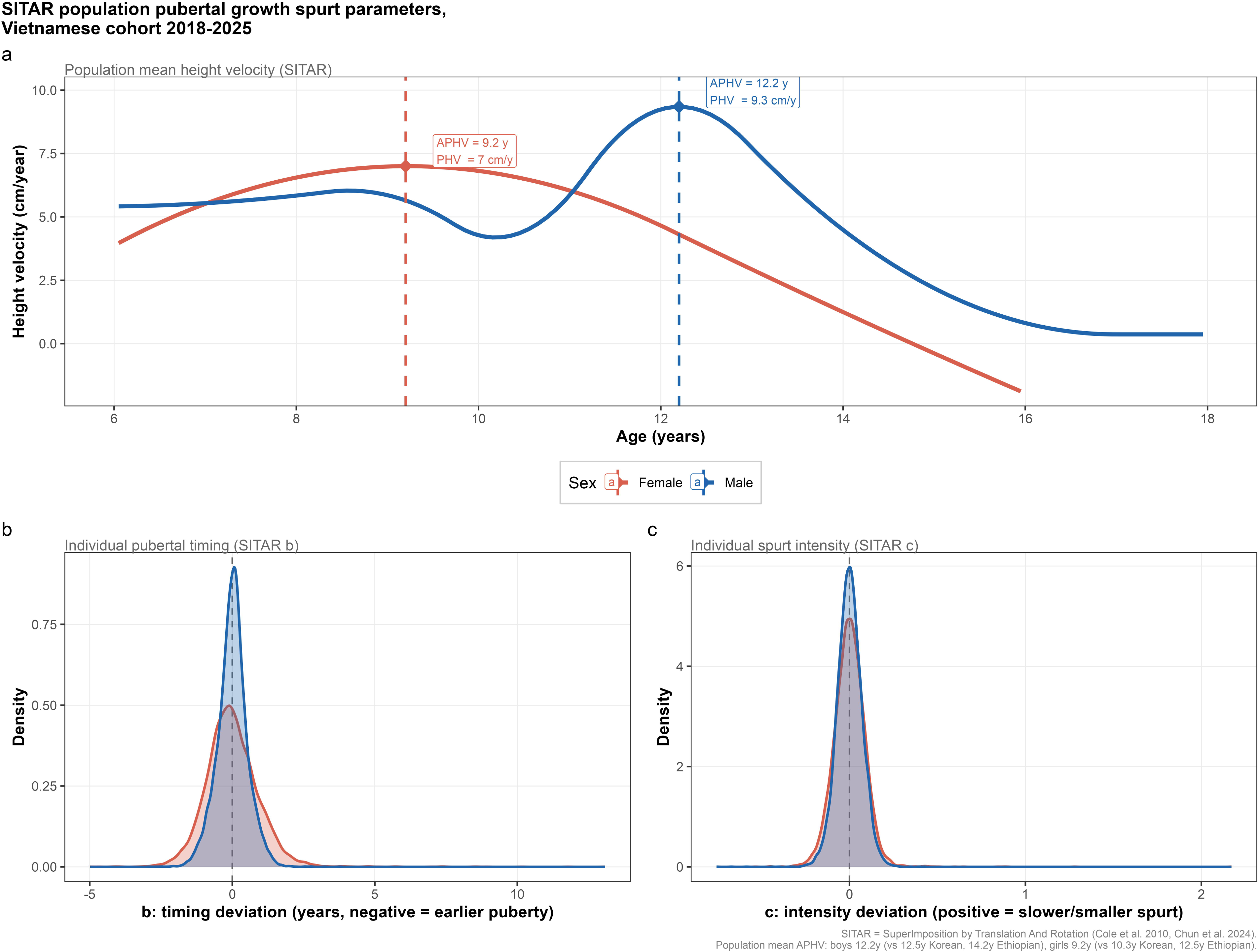
SITAR population pubertal growth spurt parameters, Vietnamese cohort 2018–2025. (a) Population mean height velocity curves by sex, derived from SITAR models fitted to longitudinal height data (boys 6 to 18 years, girls 6 to 16 years). Diamonds mark the age and magnitude of peak height velocity (APHV/PHV) for each sex. (b) Distribution of individual pubertal timing deviation (SITAR *b*) with negative values indicate earlier puberty than the population mean. (c) Distribution of individual spurt intensity deviation (SITAR *c*) with positive values indicate a slower or smaller-magnitude growth spurt. SITAR = SuperImposition by Translation And Rotation (2). Population mean APHV for boys 12.2 years (vs 12.5 years Korean (4), 14.2 years Ethiopian (12)) and for girls 9.2 years (vs 10.3 years Korean (4), 12.5 years Ethiopian (12)).

**Table 2.**
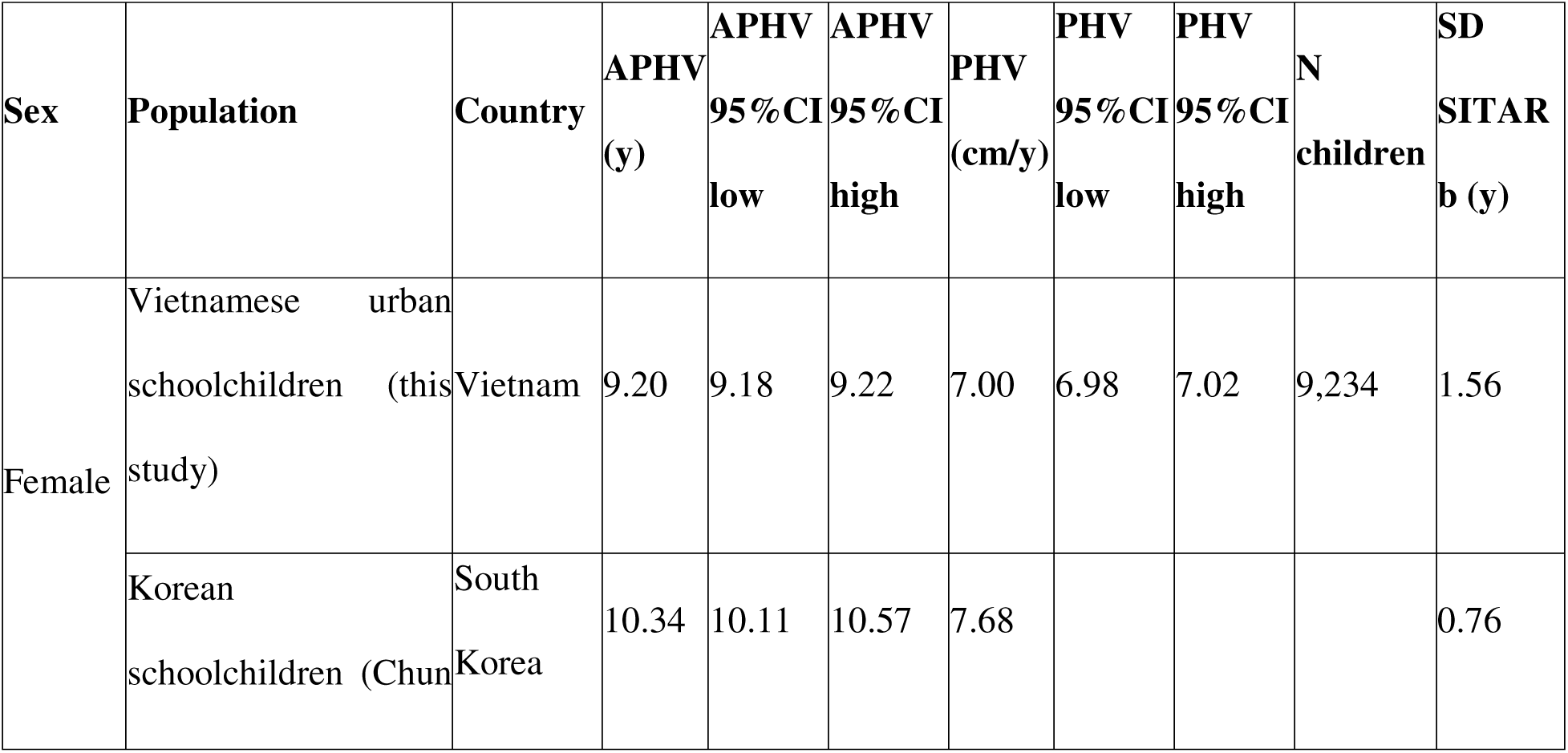
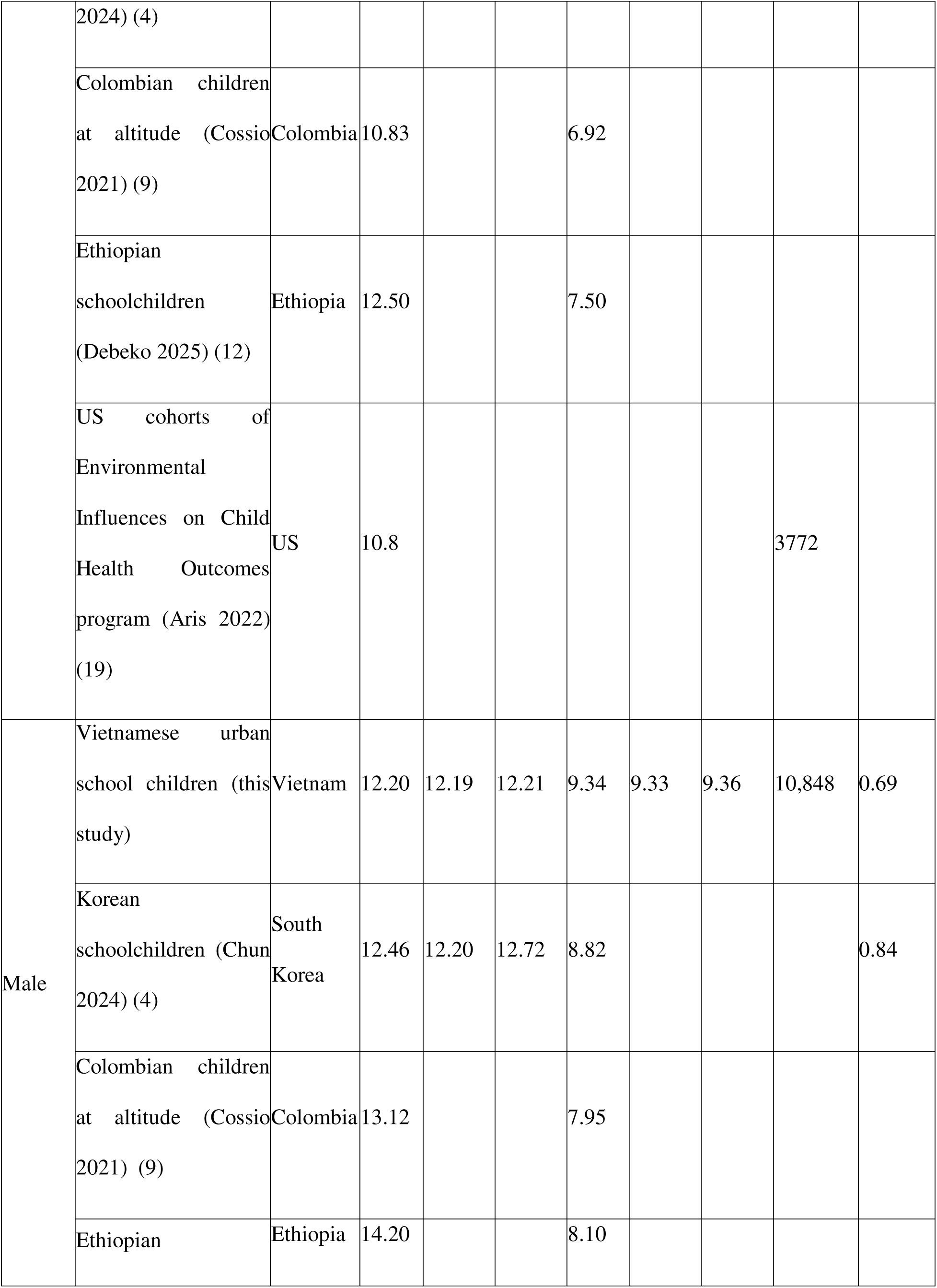
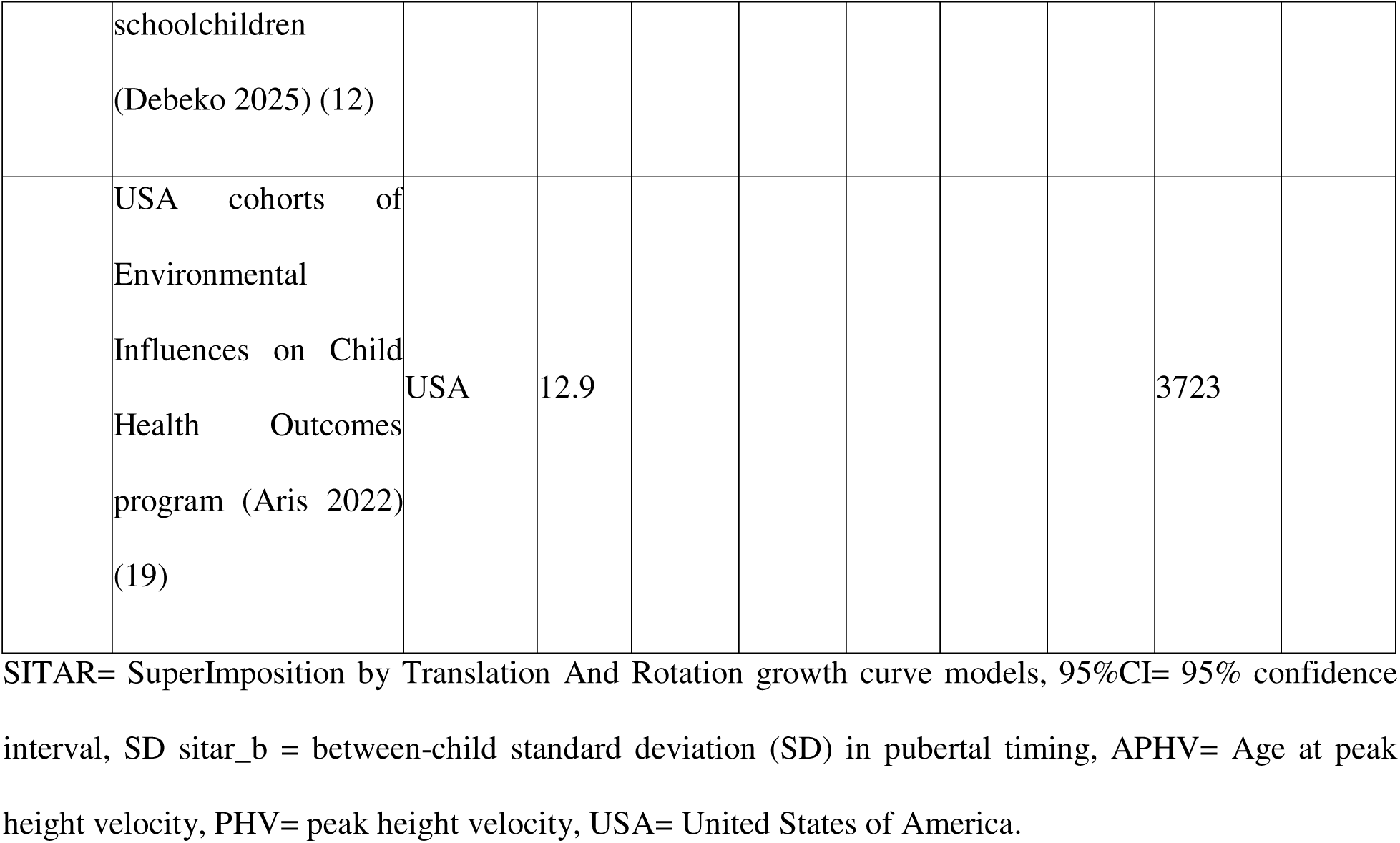
Population mean APHV and PHV (95% CI) for Vietnamese urban schoolchildren compared to published SITAR estimates.

### Individual variation in pubertal timing and intensity

Substantial inter-individual variation was observed in both the timing (SITAR b) and intensity (SITAR c) of the pubertal growth spurt (**Figure 1**). Girls showed almost twice the between-child variability in pubertal timing compared with boys (SD of b = 1.02 years in girls vs 0.52 years in boys) (**Table 1**), indicating a wider spread of individual maturational ages among girls despite their earlier population mean APHV. Individual estimated APHV (population mean adjusted for each child’s timing deviation) ranged accordingly, with a mean of 12.19 years (SD 0.52) in boys and 9.22 years (SD 1.02) in girls. Visual inspection of observed versus SITAR-fitted individual trajectories in a random subsample confirmed close correspondence between the model-fitted curves and raw longitudinal measurements across the observed age range in both sexes (**Figure 3**). BAZ trajectories during the SITAR fitting window remained relatively stable within BMI category strata. Whereas, HAZ was clearly different between BMI categories at early prepubertal age and became similar at the end pubertal age (17-18 years for boys and 15-16 years for girls) (**Figure 4**).

**Figure 2.**
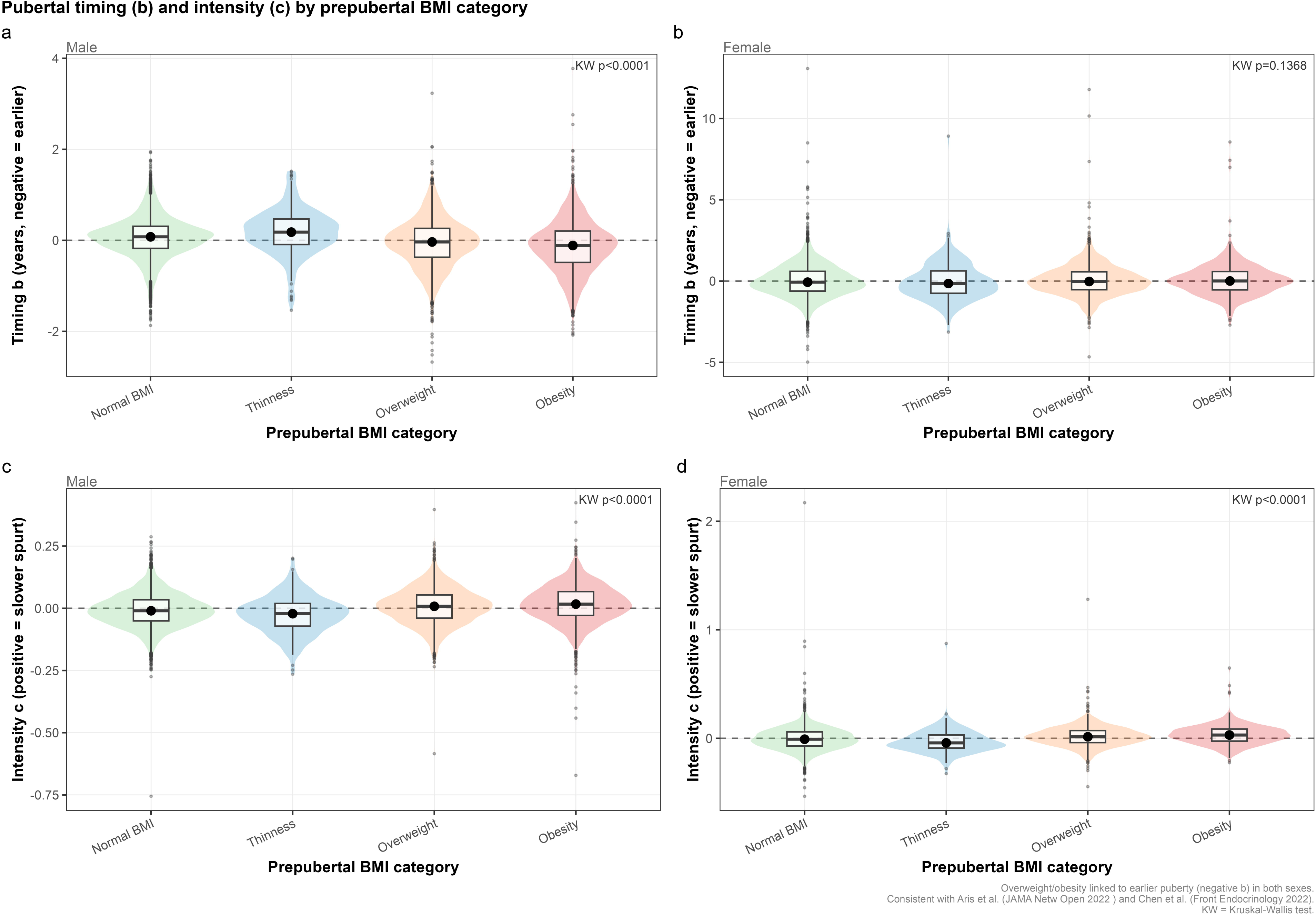
Pubertal timing (b) and intensity (c) by prepubertal BMI category. Violin and box plots of SITAR timing (b, panels a and b) and intensity (c, panels c and d) deviations across four prepubertal body mass index (BMI) categories (WHO 2007 reference), stratified by sex. Kruskal-Wallis (KW) test p-values shown for each panel. Higher BMI category was associated with significantly earlier timing in boys (panel a) but not in unadjusted comparisons in girls (panel b). Higher BMI category was associated with significantly slower/smaller spurt intensity in both sexes (panels c and d).

**Figure 3.**
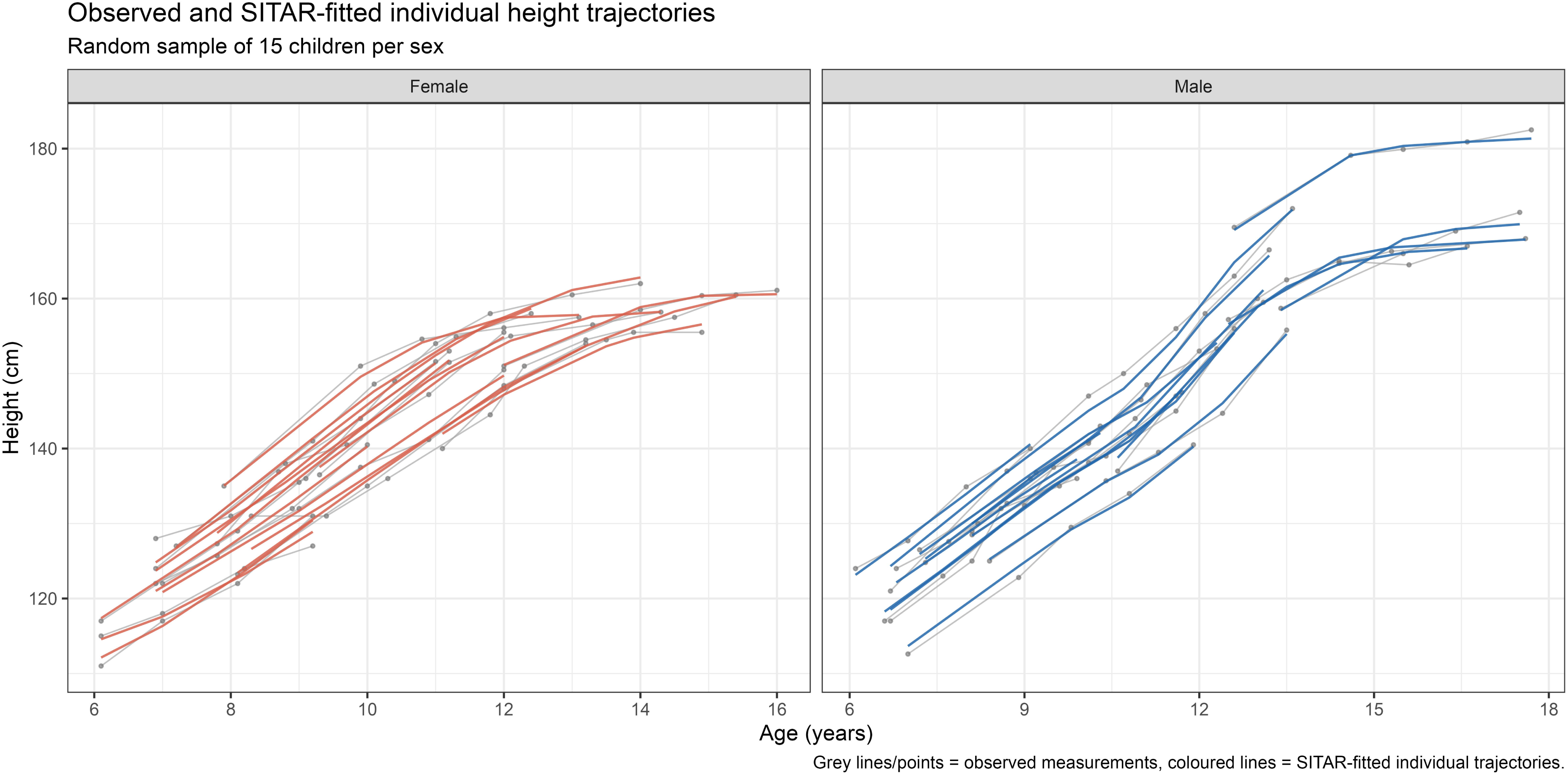
Observed and SITAR-fitted individual height trajectories. Random sample of 15 children per sex. Grey lines and points represent observed longitudinal height measurements. Colored lines represent SITAR individual fitted curves, illustrating the model’s translation-and-rotation alignment of each child’s trajectory to the population mean curve. SITAR = SuperImposition by Translation And Rotation growth curve models.

**Figure 4.**
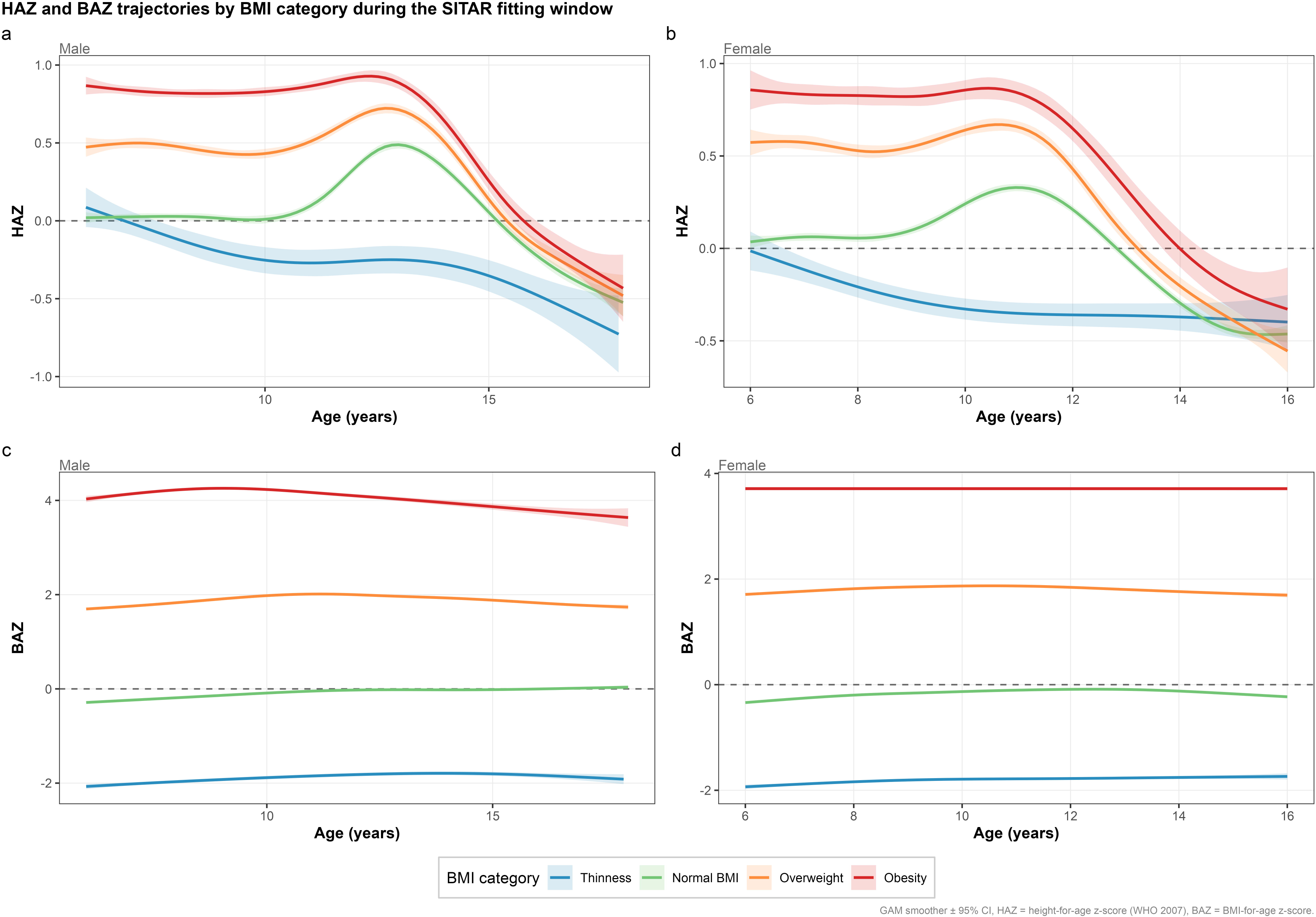
HAZ and BAZ trajectories by BMI category during the SITAR fitting window. Generalized additive model (GAM) smoothed trajectories (±95% confidence interval (CI)) of height-for-age z-score (HAZ, panels a and b) and body mass index (BMI)-for-age z-score (BAZ, panels c and d) across the SITAR fitting age range, stratified by prepubertal BMI category and sex (WHO 2007 reference). SITAR = SuperImposition by Translation And Rotation growth curve models.

### Association between prepubertal BMI status and pubertal timing and intensity

In boys, higher prepubertal BMI category was associated with earlier pubertal timing in a clear dose-dependent pattern. Mean SITAR b declined from 0.065 years in the normal BMI group to -0.136 years in the obesity group (Kruskal-Wallis p<0.0001) (**Table 3**, **Figure 2a**), corresponding to an individual estimated APHV roughly 0.2 years earlier in obese versus normal-BMI boys (12.06 vs 12.27 years). In girls, the unadjusted association between BMI category and timing was not statistically significant (p=0.137) (**Table 3**, **Figure 2b**), and the direction of the BMI-category gradient was reversed at the descriptive level, with higher BMI categories showing slightly later rather than earlier mean timing. Spurt intensity (SITAR c) showed a statistically significant gradient with BMI category in both sexes (both p<0.0001). Children with higher prepubertal BMI displayed larger positive c values, indicating a slower or smaller-magnitude growth spurt, in both boys (**Figure 2c**) and girls (**Figure 2d**).

**Table 3.**
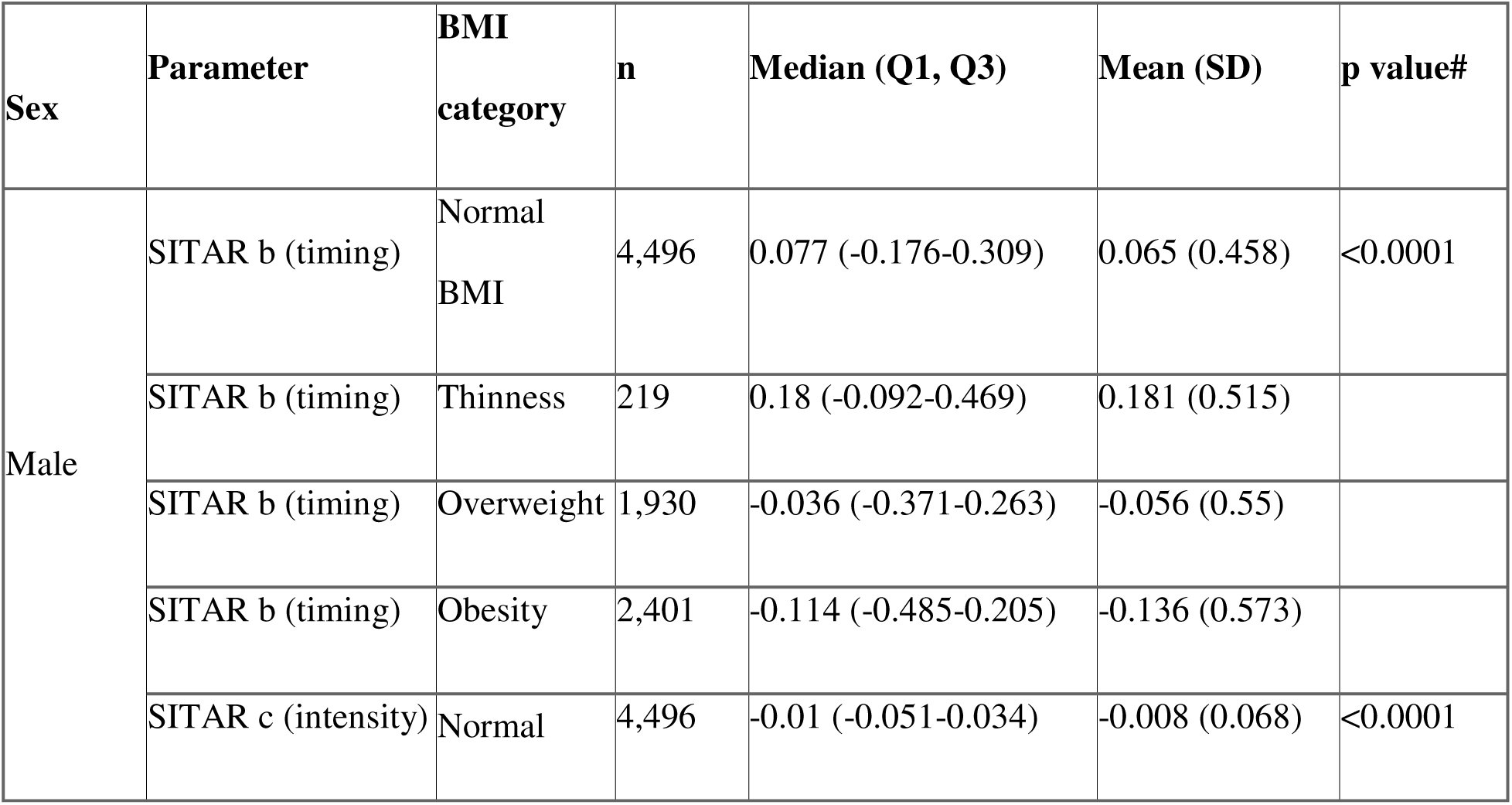
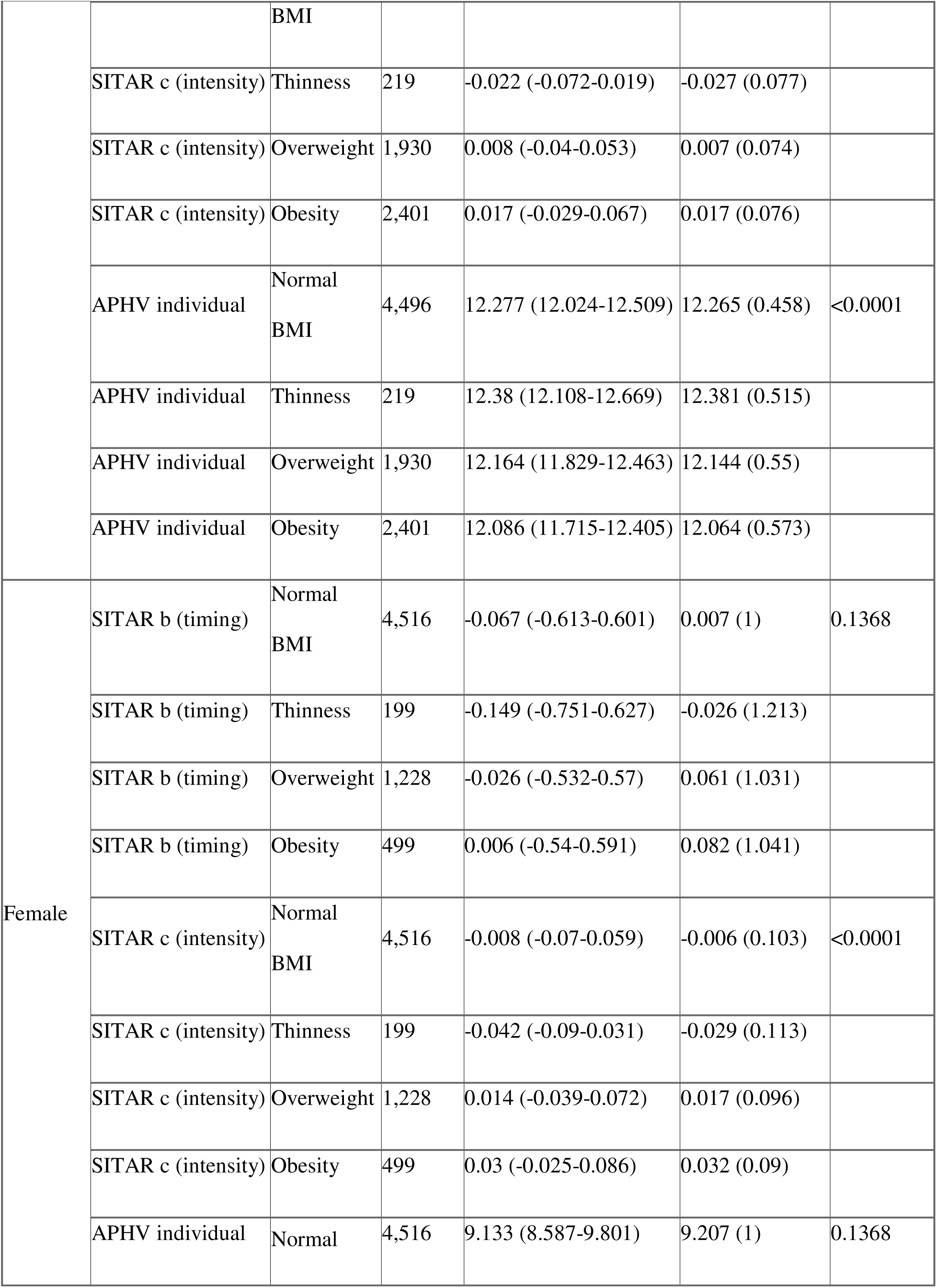
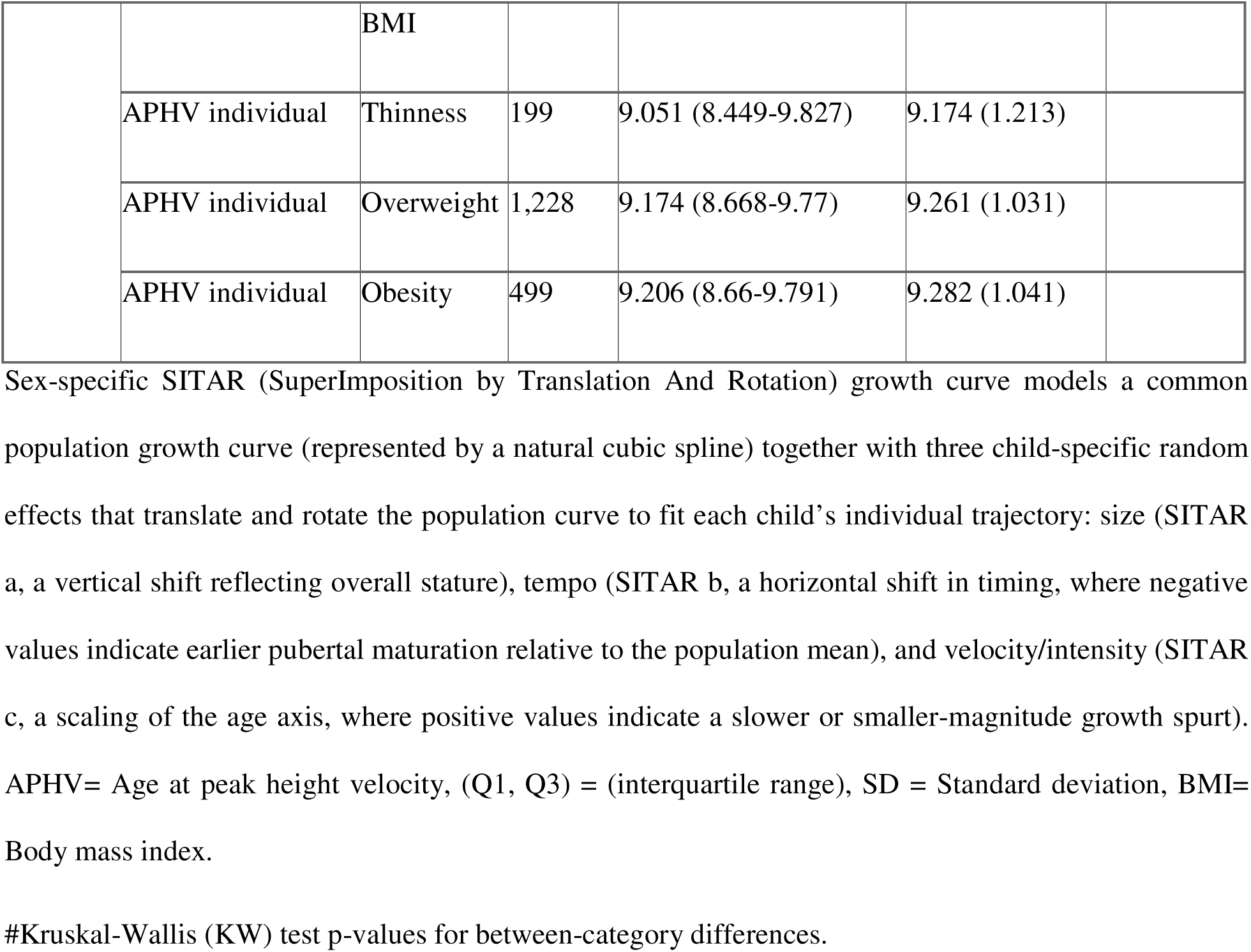
SITAR pubertal parameters (timing b, intensity c, individual APHV) by prepubertal BMI category, stratified by sex.

This apparent sex discrepancy at the univariate level for timing was resolved in the multivariable model. After adjustment for HAZ, prepubertal height velocity, BAZ trajectory slope, age at baseline, and birth year, standardized BAZ at baseline was independently associated with earlier pubertal timing in both sexes (for boys β= -0.02 per SD, 95%CI= -0.03 to -0.01, p=0.0009, for girls β= -0.09 per SD, 95%CI= - 0.14 to -0.04, p=0.0002) (**Table 4**, **Figure 7**), indicating that the null unadjusted association in girls reflected confounding by height status and growth velocity rather than a true absence of an adiposity-timing relationship. Consistent with this adjusted finding, the distribution of pubertal timing by longitudinal prepubertal BMI trajectory group showed earlier median timing in the overweight/obesity-related trajectory relative to the normal trajectory in both sexes (**Figure 8**), in agreement with prior reports linking accelerated early-life adiposity gain to earlier pubertal onset (19) and linking overweight/obesity trajectories to altered pubertal growth patterns (5).

**Table 4.** Linear mixed-effects model: standardized predictors of SITAR pubertal timing (b) and intensity (c).

| Sex | Outcome | Term | Beta (95%CI) | p value |
| --- | --- | --- | --- | --- |
| Male | SITAR b (timing) | Intercept | -0.00 (-0.06, 0.06) | 0.9900 |
|  | SITAR b (timing) | BAZ at baseline (SD) | -0.02 (-0.03, -0.01) | 0.0009 |
|  | SITAR b (timing) | HAZ at baseline (SD) | -0.17 (-0.18, -0.16) | <0.0001 |
|  | SITAR b (timing) | Prepubertal velocity (SD) | -0.18 (-0.19, -0.16) | <0.0001 |
|  | SITAR b (timing) | Delta BAZ/year (SD) | -0.02 (-0.04, 0.00) | 0.1093 |
|  | SITAR b (timing) | Age at baseline (SD) | 0.06 (0.04, 0.08) | <0.0001 |
|  | SITAR b (timing) | Birth year (SD) | 0.00 (-0.02, 0.02) | 0.6759 |
| Female | SITAR b (timing) | Intercept | -0.02 (-0.10, 0.07) | 0.6289 |
|  | SITAR b (timing) | BAZ at baseline (SD) | -0.09 (-0.14, -0.04) | 0.0002 |
|  | SITAR b (timing) | HAZ at baseline (SD) | 0.34 (0.30, 0.37) | <0.0001 |
|  | SITAR b (timing) | Prepubertal velocity (SD) | 0.11 (0.07, 0.14) | <0.0001 |
|  | SITAR b (timing) | Delta BAZ/year (SD) | 0.03 (-0.02, 0.08) | 0.2371 |
|  | SITAR b (timing) | Age at baseline (SD) | -0.19 (-0.27, -0.11) | <0.0001 |
|  | SITAR b (timing) | Birth year (SD) | -0.22 (-0.27, -0.16) | <0.0001 |
| Male | SITAR c (intensity) | Intercept | 0.00 (-0.01, 0.01) | 0.5899 |
|  | SITAR c (intensity) | BAZ at baseline (SD) | 0.00 (0.00, 0.00) | 0.0256 |
|  | SITAR c (intensity) | HAZ at baseline (SD) | 0.03 (0.03, 0.04) | <0.0001 |
|  | SITAR c (intensity) | Prepubertal velocity (SD) | 0.03 (0.03, 0.03) | <0.0001 |
|  | SITAR c (intensity) | Delta BAZ/year (SD) | 0.01 (0.00, 0.01) | <0.0001 |
|  | SITAR c (intensity) | Age at baseline (SD) | -0.01 (-0.01, -0.01) | <0.0001 |
|  | SITAR c (intensity) | Birth year (SD) | -0.01 (-0.01, -0.00) | <0.0001 |
| Female | SITAR c (intensity) | Intercept | -0.00 (-0.01, 0.01) | 0.9636 |
|  | SITAR c (intensity) | BAZ at baseline (SD) | -0.00 (-0.01, 0.00) | 0.4919 |
|  | SITAR c (intensity) | HAZ at baseline (SD) | 0.05 (0.05, 0.05) | <0.0001 |
|  | SITAR c (intensity) | Prepubertal velocity (SD) | 0.03 (0.02, 0.03) | <0.0001 |
|  | SITAR c (intensity) | Delta BAZ/year (SD) | 0.01 (0.00, 0.01) | 0.0016 |
|  | SITAR c (intensity) | Age at baseline (SD) | -0.02 (-0.03, -0.01) | <0.0001 |
|  | SITAR c (intensity) | Birth year (SD) | -0.02 (-0.03, -0.02) | <0.0001 |

City included as random intercept. All continuous predictors standardized (standard deviation (SD) units) to allow direct comparison of effect sizes. Negative beta for b = higher predictor value associated with earlier puberty, positive beta for c = larger predictor value associated with slower/smaller spurt.
SITAR= SuperImposition by Translation And Rotation growth curve models, BAZ = Body mass index (BMI) for age Z-score, HAZ = Height for age Z-score, 95%CI = 95% confidence interval.

### Geographic and secular patterns in pubertal timing

Individual estimated APHV varied modestly across the three study cities within each sex, with overlapping distributions but visibly different central tendencies on half-eye plots (**Figure 5**). Examining secular trends by birth year, individual APHV in girls declined significantly with more recent birth cohorts (β= -0.096 years per birth year, p<0.0001), indicating a shift toward earlier puberty on the order of approximately one year per decade among girls born from 2007 to 2017 (**Figure 6**). No significant secular trend was detected in boys (β= -0.004 years per birth year, p=0.117). This female-specific secular advancement in pubertal timing was corroborated in the multivariable model, where birth year remained an independent, significant predictor of earlier timing in girls (β= -0.22 per SD, 95%CI= -0.27, -0.16, p<0.0001) but not in boys (β=0.00, p=0.676) (**Table 4**).

**Figure 5.**
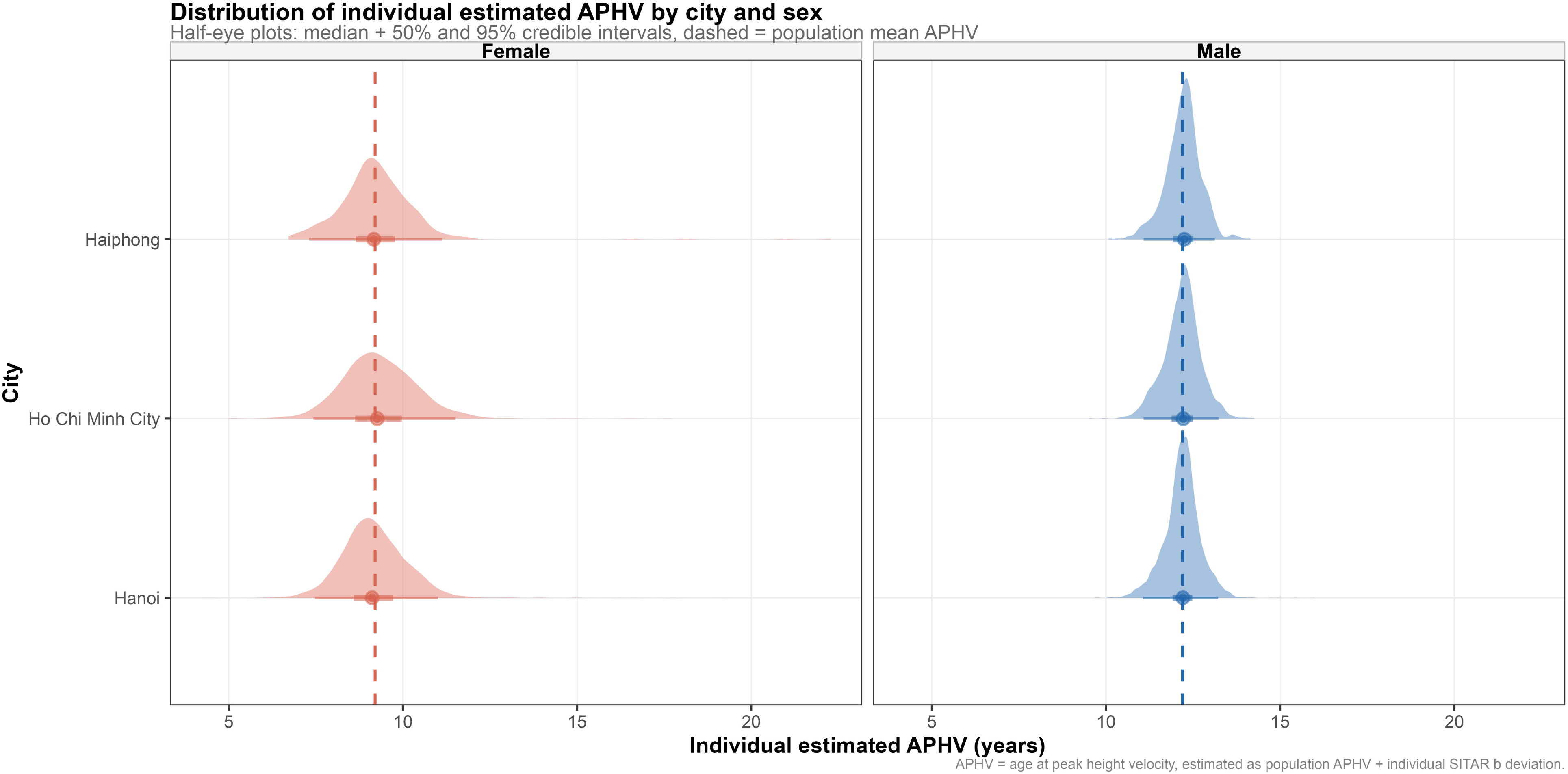
Distribution of individual estimated age at peak height velocity (APHV) by city and sex. Half-eye plots showing the median, 50% and 95% credible intervals of individual estimated APHV (population mean APHV + individual SITAR b deviation) for each of the three study cities (Hanoi, Ho Chi Minh City, Haiphong), stratified by sex. Dashed vertical lines indicate the sex-specific population mean APHV. SITAR = SuperImposition by Translation And Rotation growth curve models.

**Figure 6.**
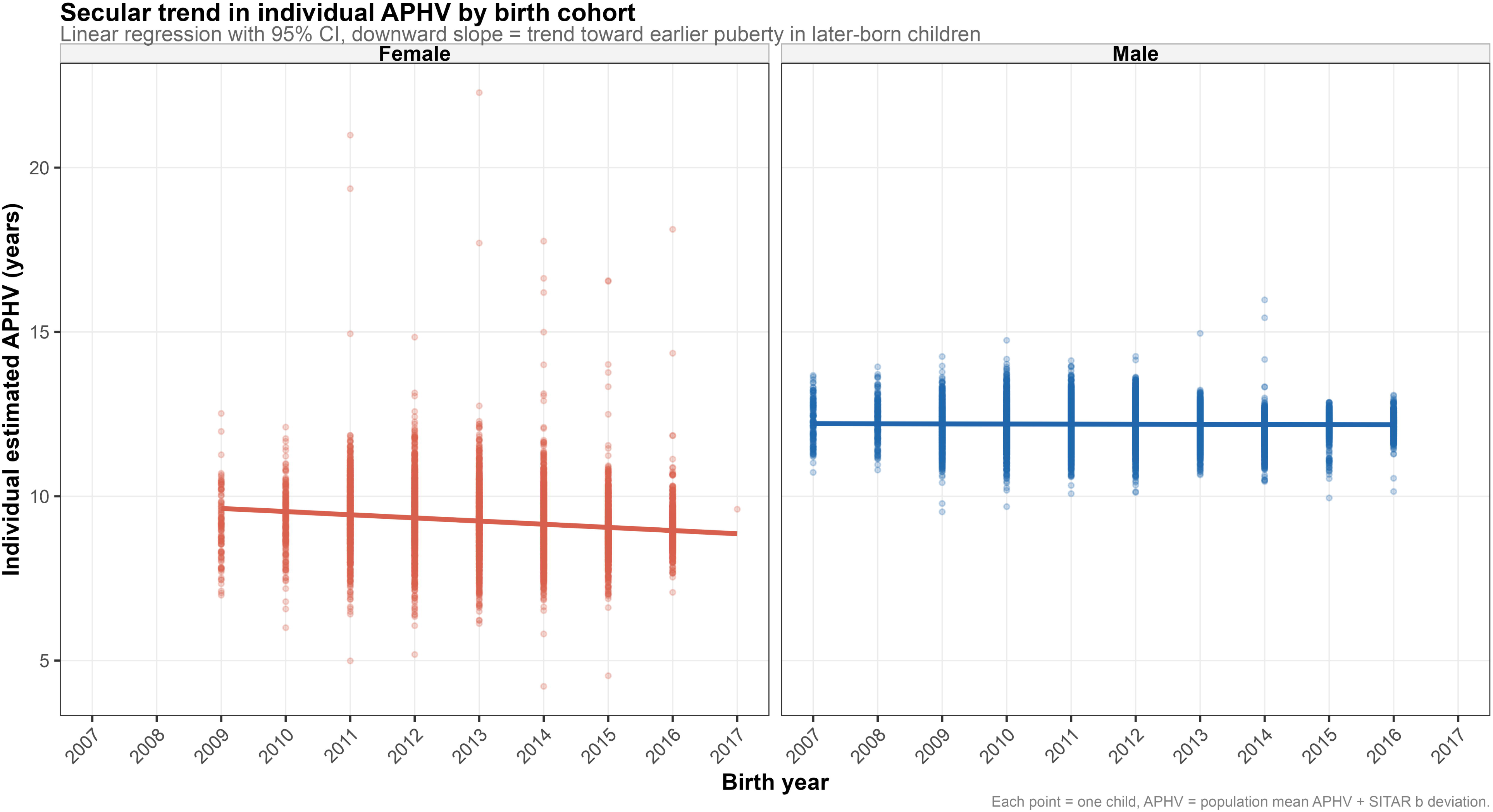
Secular trend in individual estimated age at peak height velocity (APHV) by birth cohort. Scatterplot of individual estimated APHV against birth year (2007 to 2017), with linear regression fit and 95% confidence band, stratified by sex. A significant downward trend was observed in girls (β=−0.096 years/year, p<0.0001) but not in boys (β=−0.004 years/year, p=0.117), indicating a secular shift toward earlier puberty among more recently born girls.

### Multivariable predictors of pubertal timing and intensity

In the linear mixed-effects models with city random intercept (**Table 4**, **Figure 7**), prepubertal height-for-age z-score (HAZ) and height velocity showed opposite-direction associations with pubertal timing in boys versus girls. In boys, higher HAZ and faster prepubertal height velocity were each associated with earlier timing (HAZ: β= -0.17, p<0.0001, velocity: β= -0.18, p<0.0001), whereas in girls the same predictors were associated with later timing (HAZ: β= +0.34, p<0.0001, velocity: β=+0.11, p<0.0001). In contrast, BAZ at baseline showed a consistent, same-direction association with earlier timing in both sexes, as noted above. For spurt intensity, higher HAZ, faster prepubertal velocity, and greater BAZ gain (ΔBAZ/year) were each associated with larger c values (slower/smaller spurt) in both sexes, while older age at baseline and more recent birth year were associated with smaller c values (faster/larger spurt) in both sexes (all p<0.0001) (**Table 4**).

**Figure 7.**
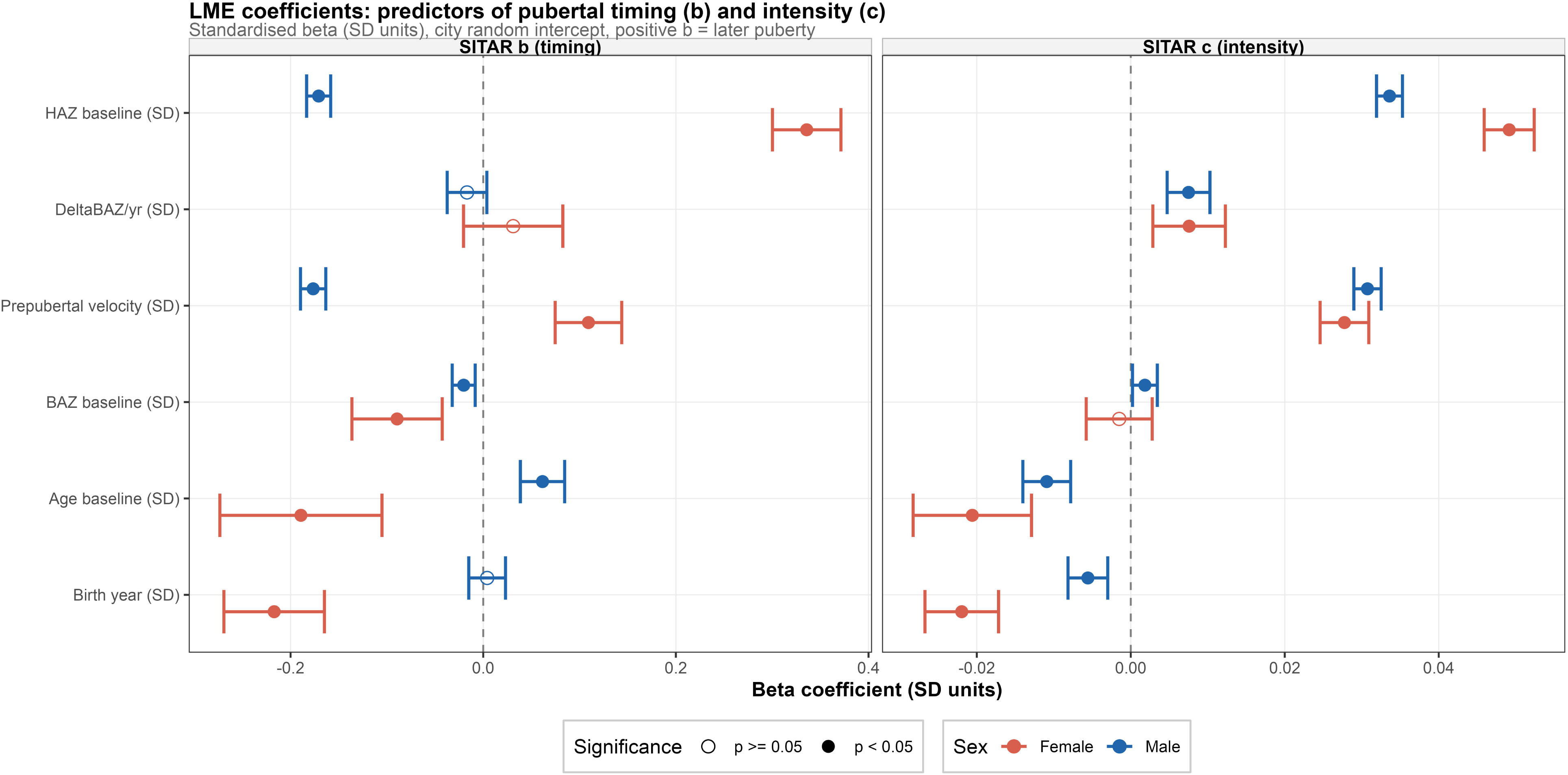
Linear mixed-effects model coefficients: predictors of pubertal timing (b) and intensity (c). Forest plot of standardized beta coefficients (standard (SD) units) from separate linear mixed-effects models for SITAR b (timing, left panel) and SITAR c (intensity, right panel), each including a city random intercept. Solid points indicate p<0.05. Positive b coefficients indicate association with later puberty. Positive c coefficients indicate association with a slower/smaller spurt. SITAR = SuperImposition by Translation And Rotation growth curve models.

**Figure 8.**
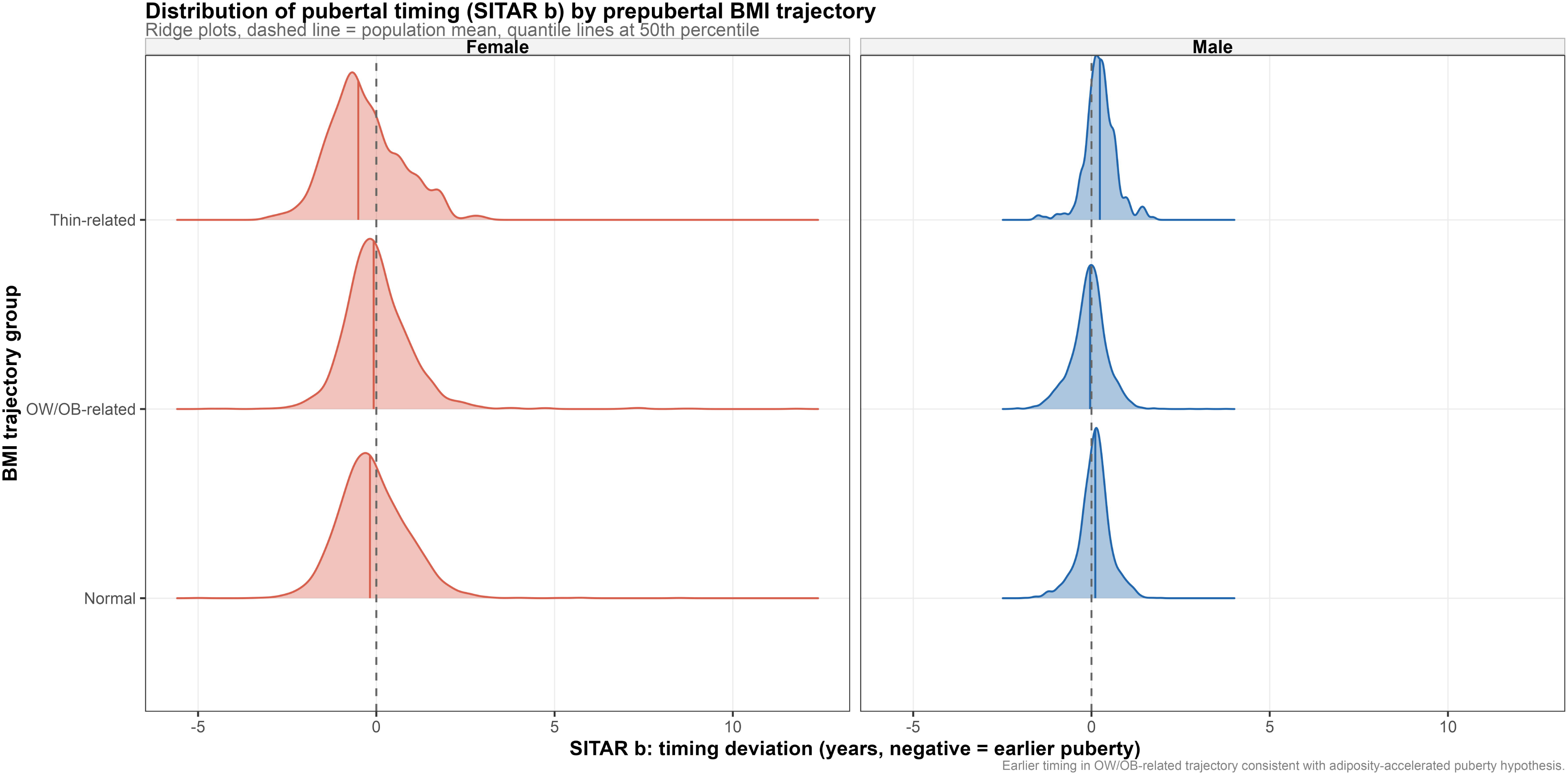
Distribution of pubertal timing (SITAR b) by prepubertal BMI trajectory. Ridge plots of SITAR b distributions across three longitudinal prepubertal body mass index (BMI) trajectory groups (Normal, overweight/obesity-related, thinness-related), stratified by sex. Dashed vertical line indicates the sex-specific population mean. Horizontal quantile lines mark the 50th percentile within each group. SITAR = SuperImposition by Translation And Rotation growth curve models.

### Prediction of early pubertal timing

Multivariable logistic regression models predicting early pubertal timing (SITAR b < 0, i.e., earlier than the population mean) achieved acceptable discrimination in boys (AUC=0.760, 95%CI= (0.748, 0.771)) and modest discrimination in girls (AUC=0.699, 95%CI= (0.682, 0.716)) (**Table 5**). In boys, higher HAZ (OR=2.38, 95%CI= (2.23, 2.55)) and faster prepubertal height velocity (OR=2.45, 95%CI= (2.26, 2.67)) were the strongest predictors of early timing, while in girls these same predictors were protective against early timing (HAZ: OR=0.58, 95%CI= (0.53, 0.63), velocity: OR=0.60, 95%CI= (0.55,0.65)), mirroring the sex-divergent pattern seen in the continuous timing model. BAZ at baseline increased the odds of early timing in both sexes (boys: OR=1.08, p=0.016 and girls: OR=1.26, p<0.0001). City of residence was retained as a significant predictor only in girls, with lower odds of early timing in Ho Chi Minh City relative to Hanoi (OR=0.67, 95%CI= (0.56, 0.80), p<0.0001) (**Table 5**).

**Table 5.** Logistic regression OR (95%CI) for early puberty.

| Sex | Term | OR (95%CI) | p value |
| --- | --- | --- | --- |
| Male | Intercept | 0.86 (0.80-0.92) | <0.0001 |
|  | BAZ baseline (SD) | 1.08 (1.01-1.14) | 0.0155 |
|  | HAZ baseline (SD) | 2.38 (2.23-2.55) | <0.0001 |
|  | Prepubertal velocity (SD) | 2.45 (2.26-2.67) | <0.0001 |
|  | Delta BAZ/year (SD) | 1.24 (1.12-1.37) | <0.0001 |
|  | Age baseline (SD) | 0.72 (0.65-0.81) | <0.0001 |
|  | Birth year (SD) | 0.84 (0.76-0.92) | 0.0002 |
|  | City: Hochiminh vs Hanoi | 1.12 (0.97-1.29) | 0.1090 |
|  | City: Haiphong vs Hanoi | 0.86 (0.71-1.05) | 0.1366 |
| Female | Intercept | 1.33 (1.17-1.51) | <0.0001 |
|  | BAZ baseline (SD) | 1.26 (1.14-1.40) | <0.0001 |
|  | HAZ baseline (SD) | 0.58 (0.53-0.63) | <0.0001 |
|  | Prepubertal velocity (SD) | 0.60 (0.55-0.65) | <0.0001 |
|  | Delta BAZ/year (SD) | 0.86 (0.76-0.96) | 0.0112 |
|  | Age baseline (SD) | 1.37 (1.14-1.65) | 0.0009 |
|  | Birth year (SD) | 1.78 (1.58-2.01) | <0.0001 |
|  | City: Hochiminh vs Hanoi | 0.67 (0.56-0.80) | <0.0001 |
|  | City: Haiphong vs Hanoi | 0.87 (0.68-1.11) | 0.2688 |

SITAR= SuperImposition by Translation And Rotation growth curve models (SITAR $b < 0$ means pubertal timing earlier than population mean). Stepwise Akaike Information Criterion (AIC) model selection applied. For male, area under the curve (AUC) =0.760, 95%CI= (0.748, 0.771) and for female, AUC=0.699, 95%CI= (0.682, 0.716). BAZ = Body mass index (BMI) for age Z-score, HAZ = Height for age Z-score, OR (95%CI) = Odds ratio (95% confidence interval).

## Discussion

This study is, to our knowledge, the first to apply the SITAR framework to characterize population and individual level pubertal growth spurt parameters in a Southeast Asian cohort. Vietnamese boys and girls reached peak height velocity earlier than previously reported Korean, Colombian, and Ethiopian cohorts modeled with the same method (4,9,12). Substantial individual heterogeneity in pubertal timing and intensity was observed, with markedly greater variability among girls than boys. Prepubertal adiposity was independently associated with earlier pubertal timing in both sexes after adjustment. Height status and growth velocity showed sex-divergent associations, and a pronounced secular trend toward earlier puberty was detected specifically in girls.

The comparatively early population APHV observed in this cohort is consistent with Vietnam’s ongoing nutritional transition, characterized by rising childhood overweight and obesity prevalence alongside improving overall nutritional status (18). Childhood adiposity is a well-established nutritional determinant of pubertal timing, plausibly acting through leptin-mediated signaling to the hypothalamic-pituitary-gonadal axis (28). Our finding that higher baseline BAZ independently predicted earlier timing in both sexes, after accounting for height status and growth velocity, is consistent with this mechanism and aligns with evidence from United States and Chinese cohorts linking early-life adiposity gain to earlier pubertal onset and altered growth patterns (5,19). Compared with Ethiopian children, who reach peak height velocity substantially later (12), the roughly two-year earlier APHV observed here likely reflects the markedly different nutritional and socioeconomic contexts of the two populations in addition to ethnic or genetic difference in maturational timing, underscoring the variation of pubertal timing owning to the childhood nutritional environment (28).

A striking and unexpected finding was the divergent direction of association between prepubertal height status and growth velocity with subsequent pubertal timing in boys versus girls in that taller, faster growing boys matured earlier, whereas taller, faster growing girls matured later. This pattern suggests that, despite evidence of broadly similar pubertal growth and skeletal maturation trajectories in boys and girls (3), the determinants of pubertal timing may not operate identically across sexes. One possible explanation is that, in girls, taller prepubertal stature and faster growth velocity partly reflect earlier initiation of pubertal growth itself under our operational definition, biasing the association toward a later apparent timing once true onset had already begun before the assigned prepubertal cutoff. Sensitivity analyses using alternative prepubertal windows would help clarify this explanation. Alternatively, the finding may reflect genuine sex-specific neuroendocrine regulation of the hypothalamic-pituitary-gonadal axis, whereby skeletal maturation and height velocity interact differently with the onset of estrogen versus androgen driven growth acceleration. This sex divergent pattern, together with recent evidence that a meaningful minority of children may not exhibit a distinct spurt at all (29,30), suggests that pubertal growth may follow more heterogeneous, population and sex specific trajectories than a single canonical model implies.

The apparent absence of a BMI-timing association in girls at the unadjusted, categorical level, contrasted with a clear and significant negative association in the multivariable model, illustrates the importance of adjusting for height status and growth velocity when examining adiposity-timing relationships, particularly in girls, where these covariates have opposing associations with timing relative to BAZ. Studies relying solely on unadjusted BMI category comparisons risk underestimating, or even reversing the apparent direction of, the true adiposity-puberty relationship in female cohorts, a methodological consideration that may explain some heterogeneity across the published literature on this association (28).

The secular advancement in pubertal timing observed among girls (approximately one year earlier per decade of more recent birth), but not boys, in this cohort substantially exceeds the roughly three-month-per-decade global average reported in a systematic review and meta-analysis of secular trends in pubertal onset (31), although that estimate was derived predominantly from high-income countries over a longer historical period. A steeper, compressed secular shift is plausible in a population undergoing rapid nutritional transition within a comparatively narrow observation window, but the relatively short birth-cohort range available in this study warrants cautious interpretation, and replication with longer follow-up is needed to confirm whether this trend persists or represents a transient feature of the cohort’s specific socioeconomic profile of urban children attending private schools.

Strengths of this study include a large, longitudinally followed cohort, use of the validated SITAR modeling framework (2), and multivariable adjustment enabling sex-specific dissection of adiposity, height status, and growth velocity effects. There are some notable limitations. First, the cohort are from a private school health system in major cities, which may not represent the broader Vietnamese pediatric population and could bias toward higher socioeconomic status. Second, the study uses anthropometric proxies for pubertal timing rather than direct clinical Tanner staging or hormonal assessment. Third, a prepubertal ascertainment window that, while chosen to precede the population mean APHV with a safety margin, may still misclassify a subset of early-maturing children, particularly girls, given their substantially greater individual variability in timing.

In brief, our findings provide the first SITAR-based characterization of pubertal growth spurt timing and intensity in Vietnamese children, reveal clinically relevant sex-specific associations with prepubertal adiposity and height status, and highlight a pronounced secular trend toward earlier puberty in girls that warrants further surveillance as Vietnam’s nutritional transition continues.

## Contributors’ statement

Nhan T. Ho did conceptualization, data curation, formal analysis, investigation, methodology, project administration, resources, software, supervision, validation, visualization, writing original draft, and writing review & editing. Quyet V. Nguyen, Anh Q. Dao, Chi T. L. Tran, and Nhan T. Ho did data curation, resources, validation. An N. Pham did data curation, resources, supervision, validation, and writing review & editing. All authors read and approved the manuscript.

## Data Sharing Statement

R codes are available from the corresponding author upon reasonable request. Individual patient-level data cannot be shared due to applicable privacy regulations and the terms of the institutional ethics approval.

## Conflict of intertest

On behalf of all authors, the corresponding author states that there is no conflict of interest

## Ethic and Consent statement

This study was approved by Vinmec Ethical Committee (approval number 0231/2024/CN/HDDD VMEC) with a waiver of individual informed consent as the study used de-identified routinely collected health data from school health examinations.

## Use of Artificial Intelligence

The authors performed all original research work regarding scientific content, analyses, interpretations and manuscript writing. The authors used AI-assisted tools for language editing and grammar checking during manuscript preparation.

